# COVID-19 vaccine effectiveness during a prison outbreak when the Omicron was the dominant circulating variant— Zambia, December 2021

**DOI:** 10.1101/2022.05.06.22274701

**Authors:** John Simwanza, Jonas Z. Hines, Danny Sinyange, Nyambe Sinyange, Chilufya Mulenga, Sarah Hanyinza, Patrick Sakubita, Nelia Langa, Haggai Nowa, Priscilla Gardner, Ngonda Saasa, Gabriel Chipeta, James Simpungwe, Warren Malambo, Busiku Hamainza, Nathan Kapata, Muzala Kapina, Kunda Musonda, Mazyanga Liwewe, Consity Mwale, Sombo Fwoloshi, Lloyd B. Mulenga, Simon Agolory, Victor Mukonka, Roma Chilengi

## Abstract

During a COVID-19 outbreak in a prison in Zambia from 14^th^ to 19^th^ December 2021, a case control study was done to measure vaccine effectiveness (VE) against infection and symptomatic infection, when the Omicron variant was the dominant circulating variant. Among 382 participants, 74.1% were fully vaccinated and the median time since full vaccination was 54 days. There were no hospitalizations or deaths. COVID-19 VE against any SARS-CoV-2 infection was 64.8% and VE against symptomatic SARS-CoV-2 infection was 72.9%. COVID-19 vaccination helped protect incarcerated persons against SARS-CoV-2 infection during an outbreak while Omicron was the dominant variant in Zambia.

## Introduction

COVID-19 continues to ravage the globe as new SARS-CoV-2 variants emerge and cause new waves of infections. The highly transmissible Omicron variant emerged in late 2021, rapidly spreading to many countries and leading to record-breaking case counts globally. In early December 2021, Zambia recorded a rapid rise in confirmed COVID-19 cases, coinciding with confirmation of the Omicron variant in the country. Zambia recorded a daily average of 1,422 confirmed COVID-19 cases in December 2021, compared to an average 15 cases daily the previous month, in November 2021. From December 4^th^ to 24^th^, 2021, all sequenced specimens in Zambia were of the Omicron variant (1).

Congregate living settings, including prisons, are at high risk for COVID-19 outbreaks (2). Congested living conditions make physical distancing and avoidance of crowds challenging in many prison settings. Although difficult to enforce, use of face masks in high SARS-CoV-2 transmission risk settings can limit the rapid spread of infection (3).

On December 14^th^, 2021, after not recording any COVID-19 cases for months, a local prison in Lusaka noted an increase in the number of residents with respiratory symptoms, 19 of whom tested positive for COVID-19 with rapid diagnostic tests (RDTs). The prison notified the Lusaka District Health Office on the same day and an outbreak investigation was initiated. Mitigation measures were reinforced, including mask distribution and mandatory masking, cohorting persons testing positive and provision of hand hygiene stations throughout the prison. Of 767 persons incarcerated at the facility during the outbreak, 241 (31.4%) tested positive for COVID-19 from December 14^th^ to 19^th^, 2021.

COVID-19 vaccines have shown remarkable effectiveness, particularly for reducing COVID-19 severity (4). COVID-19 vaccine effectiveness (VE) against the Omicron variant is lower than against other variants (5,6), although data from the African continent are limited (7–9). Correctional services in Zambia achieved high COVID-19 vaccine coverage among incarcerated persons prior to the Omicron variant wave through onsite vaccination clinics and enhanced COVID-19 screening and testing with encouragement of vaccination for those testing negative. Of the incarcerated persons at this local prison, 619 (80.7%) were vaccinated when the outbreak occurred (compared to 12.6% of the eligible general population in Zambian at the time). We measured VE among incarcerated persons during this outbreak in Zambia.

## Methods

A case-control study of SARS-CoV-2 infection and symptomatic infection by vaccination status was conducted among persons incarcerated at the prison facility during the outbreak from 15^th^ to 20^th^ December 2021. The facility is in Lusaka, the national capital of Zambia, and houses men aged ≥13 years awaiting court cases or transfer to other prisons after sentencing. The prison has five cells built to house 92 persons, although at the outbreak onset 767 persons were incarcerated (i.e., > 800% beyond capacity).

During the outbreak, all incarcerated persons were tested for COVID-19 using Panbio COVID-19 Ag rapid test (Abbott Rapid Diagnostics Jena GmbH, Germany) from December 14^th^ to 19^th^, 2021. Cases and controls were incarcerated persons testing COVID-19 positive and negative, respectively. Participants were recruited into the study between December 20^th^ to 24^th^, 2021 by reviewing the outbreak line list maintained by the prison health team and district health office, attempting to frequency match controls by cases’ cell assignment. Verbal consent was obtained from the participants, and for minors aged 13–17 years consent was obtained from the prison warden as per Zambia Correctional Services policy. This study was approved by the ERES Ethics review board and National Health Research Authority. The activity was reviewed by US Centers for Disease Control and Prevention and was conducted consistent with applicable Federal law and CDC policy.

A standardized questionnaire that included information on demographics and medical history, including COVID-19 vaccination and test results was administered by trained interviewers. Self-reported COVID-19 test results were confirmed with the line list where the RDT results were recorded. Vaccination status was cross-referenced with the national registry.

Full vaccination was defined as having received the 1^st^ dose of a one-dose vaccine or 2^nd^ dose of a two-dose vaccine ≥14 days before COVID-19 testing. Partial vaccination was defined as having received the 1^st^ dose of a two-dose vaccine ≥14 days before COVID-19 testing but not the 2^nd^ dose yet or receiving the 2^nd^ dose ≤13 days before testing. Persons who were partially vaccinated or indeterminate status (i.e., received their 1^st^ dose of the One dose vaccine 0-13 days before testing, or had an unknown type of COVID-19 vaccine) were excluded from analyses. Multivariable logistic regression was used to calculate the odds of SARS-CoV-2 infection and symptomatic infection, adjusting for age, number of comorbidities, jail cell, and self-reported mask use. VE was calculated as 1 minus the adjusted odds ratio times 100. To reduce the risk of misclassification bias related to false negative RDT test results, we conducted a sensitivity analysis excluding symptomatic controls.

## Results

In total, 385 (50.2%) of the 767 incarcerated persons present during the outbreak were reached for interview and 382 (49.8%) consented to being enrolled in the study. All were males, with a median age of 28 years (interquartile range [IQR]: 21-36 years) (Table 1). Overall, 84 (22.0%) had at least one comorbidity, with HIV being most common (n = 40, 10.5%). Only 16 (4.1%) reported having had COVID-19 previously.

**Table 1.**
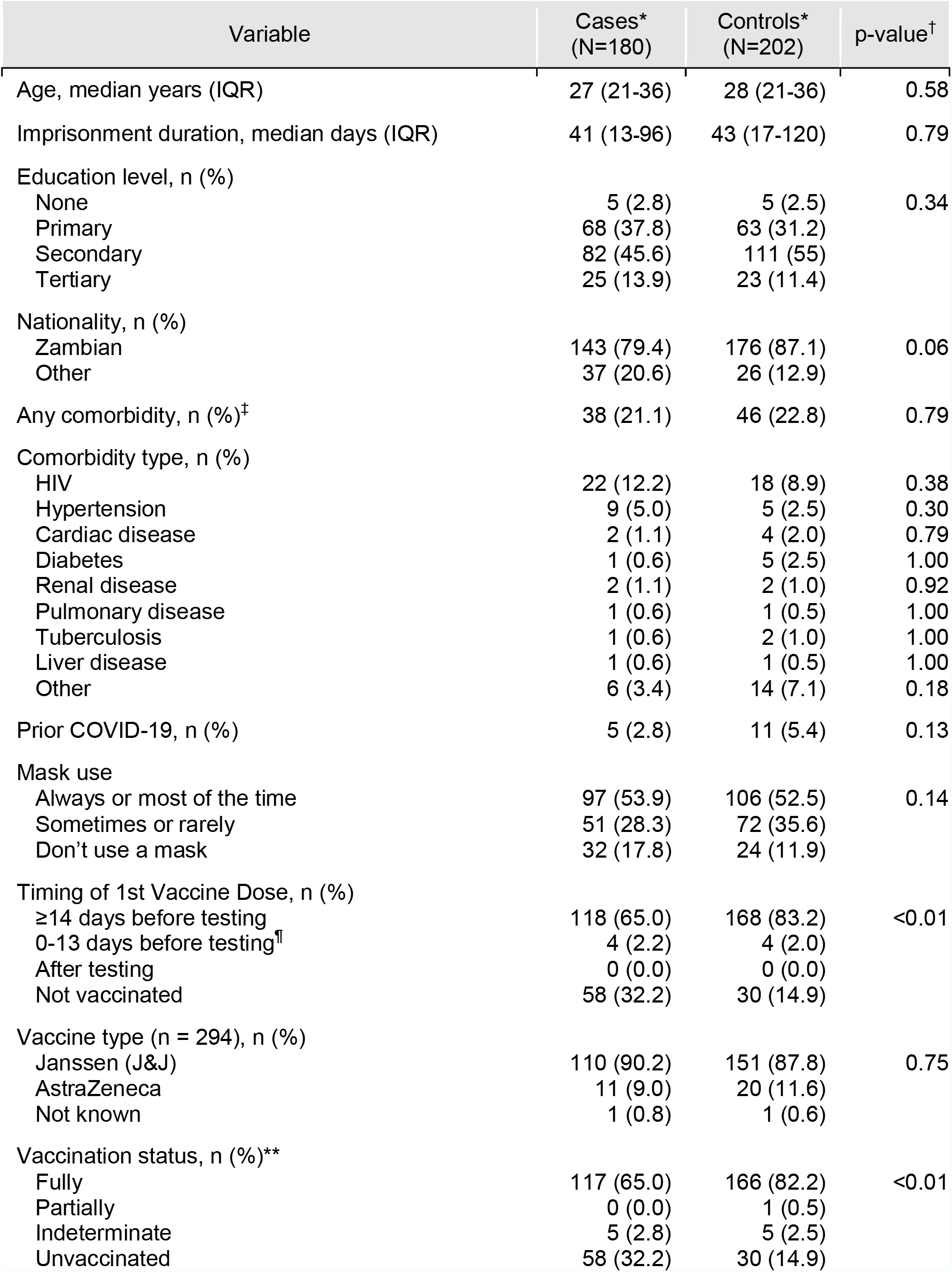
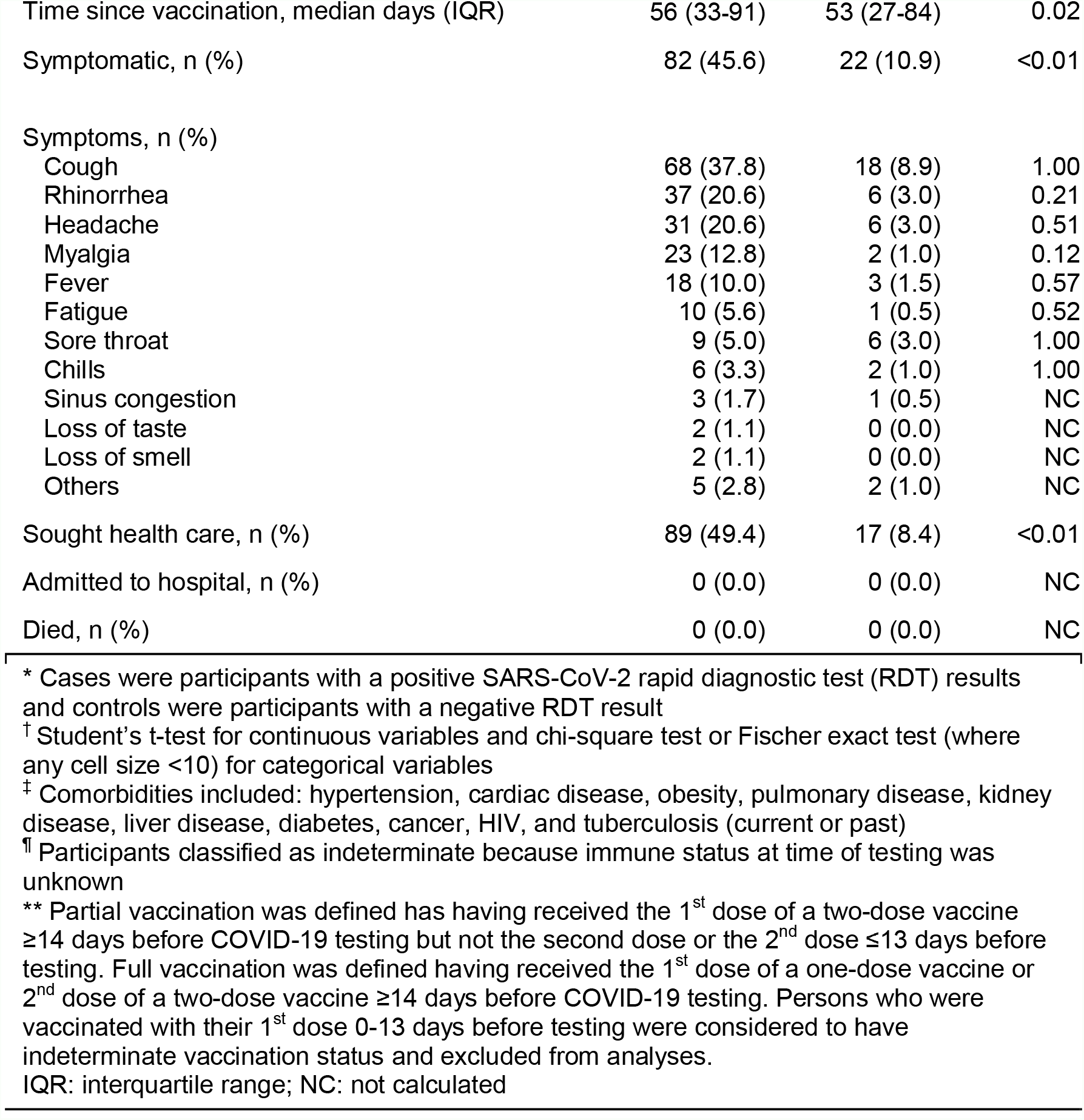
Characteristics and outcomes of cases and controls during an COVID-19 outbreak in a prison in Zambia, December 2021

| Variable | Cases*<br>(N=180) | Controls*<br>(N=202) | p-value<sup>†</sup> |
| --- | --- | --- | --- |
| Age, median years (IQR) | 27 (21-36) | 28 (21-36) | 0.58 |
| Imprisonment duration, median days (IQR) | 41 (13-96) | 43 (17-120) | 0.79 |
| Education level, n (%) |  |  |  |
| None | 5 (2.8) | 5 (2.5) | 0.34 |
| Primary | 68 (37.8) | 63 (31.2) |  |
| Secondary | 82 (45.6) | 111 (55) |  |
| Tertiary | 25 (13.9) | 23 (11.4) |  |
| Nationality, n (%) |  |  |  |
| Zambian | 143 (79.4) | 176 (87.1) | 0.06 |
| Other | 37 (20.6) | 26 (12.9) |  |
| Any comorbidity, n (%)<sup>‡</sup> | 38 (21.1) | 46 (22.8) | 0.79 |
| Comorbidity type, n (%) |  |  |  |
| HIV | 22 (12.2) | 18 (8.9) | 0.38 |
| Hypertension | 9 (5.0) | 5 (2.5) | 0.30 |
| Cardiac disease | 2 (1.1) | 4 (2.0) | 0.79 |
| Diabetes | 1 (0.6) | 5 (2.5) | 1.00 |
| Renal disease | 2 (1.1) | 2 (1.0) | 0.92 |
| Pulmonary disease | 1 (0.6) | 1 (0.5) | 1.00 |
| Tuberculosis | 1 (0.6) | 2 (1.0) | 1.00 |
| Liver disease | 1 (0.6) | 1 (0.5) | 1.00 |
| Other | 6 (3.4) | 14 (7.1) | 0.18 |
| Prior COVID-19, n (%) | 5 (2.8) | 11 (5.4) | 0.13 |
| Mask use |  |  |  |
| Always or most of the time | 97 (53.9) | 106 (52.5) | 0.14 |
| Sometimes or rarely | 51 (28.3) | 72 (35.6) |  |
| Don't use a mask | 32 (17.8) | 24 (11.9) |  |
| Timing of 1st Vaccine Dose, n (%) |  |  |  |
| ≥14 days before testing | 118 (65.0) | 168 (83.2) | <0.01 |
| 0-13 days before testing<sup>¶</sup> | 4 (2.2) | 4 (2.0) |  |
| After testing | 0 (0.0) | 0 (0.0) |  |
| Not vaccinated | 58 (32.2) | 30 (14.9) |  |
| Vaccine type (n = 294), n (%) |  |  |  |
| Janssen (J&J) | 110 (90.2) | 151 (87.8) | 0.75 |
| AstraZeneca | 11 (9.0) | 20 (11.6) |  |
| Not known | 1 (0.8) | 1 (0.6) |  |
| Vaccination status, n (%)<sup>**</sup> |  |  |  |
| Fully | 117 (65.0) | 166 (82.2) | <0.01 |
| Partially | 0 (0.0) | 1 (0.5) |  |
| Indeterminate | 5 (2.8) | 5 (2.5) |  |
| Unvaccinated | 58 (32.2) | 30 (14.9) |  |
| Time since vaccination, median days (IQR) | 56 (33-91) | 53 (27-84) | 0.02 |
| Symptomatic, n (%) | 82 (45.6) | 22 (10.9) | <0.01 |
| Symptoms, n (%) |  |  |  |
| Cough | 68 (37.8) | 18 (8.9) | 1.00 |
| Rhinorrhea | 37 (20.6) | 6 (3.0) | 0.21 |
| Headache | 31 (20.6) | 6 (3.0) | 0.51 |
| Myalgia | 23 (12.8) | 2 (1.0) | 0.12 |
| Fever | 18 (10.0) | 3 (1.5) | 0.57 |
| Fatigue | 10 (5.6) | 1 (0.5) | 0.52 |
| Sore throat | 9 (5.0) | 6 (3.0) | 1.00 |
| Chills | 6 (3.3) | 2 (1.0) | 1.00 |
| Sinus congestion | 3 (1.7) | 1 (0.5) | NC |
| Loss of taste | 2 (1.1) | 0 (0.0) | NC |
| Loss of smell | 2 (1.1) | 0 (0.0) | NC |
| Others | 5 (2.8) | 2 (1.0) | NC |
| Sought health care, n (%) | 89 (49.4) | 17 (8.4) | <0.01 |
| Admitted to hospital, n (%) | 0 (0.0) | 0 (0.0) | NC |
| Died, n (%) | 0 (0.0) | 0 (0.0) | NC |

\* Cases were participants with a positive SARS-CoV-2 rapid diagnostic test (RDT) results and controls were participants with a negative RDT result
† Student's t-test for continuous variables and chi-square test or Fischer exact test (where any cell size <10) for categorical variables
‡ Comorbidities included: hypertension, cardiac disease, obesity, pulmonary disease, kidney disease, liver disease, diabetes, cancer, HIV, and tuberculosis (current or past)
¶ Participants classified as indeterminate because immune status at time of testing was unknown
\*\* Partial vaccination was defined as having received the 1<sup>st</sup> dose of a two-dose vaccine ≥14 days before COVID-19 testing but not the second dose or the 2<sup>nd</sup> dose ≤13 days before testing. Full vaccination was defined as having received the 1<sup>st</sup> dose of a one-dose vaccine or 2<sup>nd</sup> dose of a two-dose vaccine ≥14 days before COVID-19 testing. Persons who were vaccinated with their 1<sup>st</sup> dose 0-13 days before testing were considered to have indeterminate vaccination status and excluded from analyses.
IQR: interquartile range; NC: not calculated

Overall, 294 (77.0%) participants received ≥1 COVID-19 vaccine dose; of these 283 (96.3%) were fully vaccinated, 1 (0.3%) partially vaccinated and 10 (3.4%) indeterminate. Among fully vaccinated participants, 253 (89.4%) received the one-dose Janssen vaccine and 30 (10.6%) received the two-dose AstraZeneca vaccines. None had received an additional (‘booster’) vaccine dose. The median time since receipt of a full primary vaccine series was 54 days (IQR: 28-85 days).

There were 180 (47.1%) COVID-19 positive incarcerated persons (i.e., cases) and 202 (52.9%) COVID-19 negative persons (i.e., controls). Among positive cases, 117 (65.0%) were in fully vaccinated persons (i.e., breakthrough infections). Of the 16 (4.1%) persons reporting prior confirmed COVID-19, 5 (29.4%) were positive during the outbreak (i.e., reinfections).

Overall, 104 (27.2%) persons reported any COVID-19 symptoms, with a greater proportion among cases (45.6% vs. 10.9%; p <0.01, chi-square test). The most common symptoms among COVID-19 cases were cough (37.8%), rhinorrhoea (20.6%), headache (17.2%), and myalgias (12.8%) (Table 1). Eighty-nine (49.4%) of the 180 participants with cases sought medical care at the prison clinic, and none (0%) were admitted or died. Cases were more likely to have sought health care (49.4% vs. 8.4%, p <0.01, chi-square test). There were no differences among the cases and controls in terms of age, number of comorbidities, prior Covid-19, or mask use.

Cases were less likely to be fully vaccinated than controls (65.0% versus 82.2%; p<0.01, chi-square test) (Table 2). VE against SARS-CoV-2 infection was 64.8% (95% confidence interval [CI]: 36.1-81.0%) and VE against symptomatic SARS-CoV2 infection was 72.9% (95% CI: 42.0-87.5%). VE was higher for those vaccinated within the past 60 days compared to >60 days before COVID-19 testing, although the confidence intervals overlapped (VE for SARS-CoV-2 infection: 74.6% [95% CI: 50.3-87.4%] vs. 54.2% [95% CI: 6.9-77.9%], respectively). VE of Janssen vaccine against SARS-CoV-2 infection was 63.6% (95% CI: 33.6-80.5%) and against symptomatic infection was 73.0% (95% CI: 41.6-87.7%). VE of AstraZeneca against SARS-CoV-2 infection was 89.4% (95% CI: 59.5-97.8%) and against symptomatic infection was 85.1% (95% CI: 19.5-98.0%). The sensitivity analysis excluding symptomatic controls did not meaningfully change the VE estimates (VE against SARS-CoV-2 infection was 64.1% [95% CI: 35.8- 80.2%] and VE against symptomatic SARS-CoV2 infection was 73.4% [95% CI: 44.6-87.4%]). Forty persons (10.5%) in the analysis reported being HIV positive, among whom 77.3% of cases were vaccinated compared to 94.4% of controls (p = 0.20, chi-square test). Among PLHIV, VE against SARS-CoV-2 infection was 82.2% (95% CI: -107.0-99.4%).

**Table 2.** COVID-19 vaccine effectiveness against Omicron variant – Zambia, December 2021

| Vaccination status | Fully vaccinated, n (%) |  | Vaccine effectiveness, % (95% CI)* |  |
| --- | --- | --- | --- | --- |
|  | Cases | Controls | SARS-CoV-2 infection | Symptomatic infection |
| Vaccine brand<sup>†</sup> |  |  |  |  |
| Any vaccine (n = 371) | 117 (66.9) | 166 (84.7) | 64.8 (36.1-81.0) | 72.9 (42.0-87.5) |
| Janssen (n = 341) | 106 (64.6) | 147 (83.1) | 63.6 (33.6-80.5) | 73.0 (41.6-87.7) |
| AstraZeneca (n = 118) | 11 (15.9) | 19 (38.8) | 89.4 (59.5-97.8) | 85.1 (19.5-98.0) |
| Vaccination timing<sup>†</sup> |  |  |  |  |
| ≤60 days (n = 249) | 59 (50.4) | 102 (77.3) | 74.6 (50.3-87.4) | 83.7 (60.8-93.6) |
| >60 days (n = 210) | 58 (50.0) | 64 (68.1) | 54.2 (6.9-77.9) | 53.2 (-16.6-81.6) |

\* Adjusted for age, number of comorbidities, jail cell, and mask use
<sup>†</sup> Analyses presented for fully vaccinated persons only. One person was partially vaccinated and 10 were indeterminate vaccination status (i.e., vaccinated with their 1<sup>st</sup> dose 0-13 days before COVID-19 testing)
CI: confidence interval

## Discussion

During a prison COVID-19 outbreak in Zambia, COVID-19 vaccination protected against SARS-CoV-2 infection and symptomatic illness while Omicron was the dominant strain in the country (1). These findings provide important evidence that might help increase COVID-19 vaccination in Zambia, where many more Zambians need to be vaccinated to reach the African Union targets adopted by the country (10).

The SARS-CoV-2 Omicron variant contains numerous mutations to the spike protein which may lead to immune evasion. VE estimates reported here are lower compared to prior strains (11,12), and the high proportion of breakthrough infections in this outbreak supports in theory the possibility of immune evading capability of Omicron. In this study, VE was higher for symptomatic illness than any infection; VE for severe illness could not be accessed because no persons were hospitalized or died during this outbreak although other studies have demonstrated durability against this important outcome (13). Similarly, VE of a booster dose could not be assessed because Zambia did not begin offering a booster dose until January 2022. The relatively short time since vaccination might explain the relatively higher effectiveness findings against Omicron (6,14,15). However, there was a suggestion of waning immunity when comparing the VE point estimates of participants fully vaccinated over two months before the outbreak with those vaccinated within the past two months.These findings are encouraging considering alarm raised early in the Omicron wave about the vaccines widely available in Africa being ineffective against reducing SARS-CoV-2 transmission (16).

Notably, symptomology was mild among participants, which is consistent with reports from other countries that experienced Omicron surges (9,17). Animal evidence suggests this might be related to tropism of Omicron for upper respiratory epithelium relative to the lower respiratory tract (18). This finding might also reflect the relatively younger age of persons in this outbreak. The role of pre-existing immunity in Zambia from natural infection remains uncertain and serosurveys would be useful to understand to what proportion of people in Zambia have been infected with SARS-CoV-2.

This study had several limitations. All cases were confirmed with RDTs, which have lower sensitivity than PCR tests (19). Additionally, beyond initial test results, serial testing was not available meaning some controls might have been in their incubation period at the time of testing and therefore misclassified. Omicron was not confirmed by genomic sequencing in this outbreak, meaning the outbreak could have been from another strain; however, Omicron was already dominant in Zambia before this outbreak occurred (20). Lastly, although few participants reported a prior confirmed SARS-CoV-2 infection, the actual number might be much higher considering only a small proportion of cases are confirmed in Zambia (21).

Rapid investigation of an outbreak in a closed setting demonstrated VE of COVID-19 vaccines against Omicron infection in Zambia. COVID-19 vaccination remains a critical tool in decreasing SARS-CoV-2 transmission and severity especially when coupled with a layered prevention including well-fitting facemask use, hand hygiene, limiting large gatherings, and adequate ventilation and/or outdoor gatherings. Continuing to rapidly scale COVID-19 vaccination to all eligible persons in Zambia can help prevent SARS-CoV-2 transmission and symptomatic COVID-19.

### Attribution of Support

This work has been supported by the President’s Emergency Plan for AIDS Relief (PEPFAR) through the Centers for Disease Control and Prevention (CDC) and the CDC Emergency Response to the COVID-19 pandemic.

### Disclaimer

The findings and conclusions in this report are those of the author(s) and do not necessarily represent the official position of the funding agencies. Use of trade names is for identification only and does not imply endorsement by the funding agencies.

## Data Availability

All data produced in the present work are contained in the manuscript

